# Detection of anti–SARS-CoV-2 mucosal IgA in clinical saliva samples after a dose of Novavax COVID-19 vaccine

**DOI:** 10.1101/2025.01.21.25320906

**Authors:** Mingzhu Zhu, Edmond Massuda, Urvashi Patel, Gordon Chau, Raj Kalkeri, Shane Cloney-Clark, Katherine Smith, Susan Neal, Joyce S. Plested, Raburn M. Mallory, Chijioke Bennett

**Affiliations:** Novavax, Inc., 700 Quince Orchard Road, Gaithersburg, Maryland, 20878 USA

**Keywords:** immunogenicity, booster, respiratory, virus, IgG, IgA, mucosal

## Abstract

Immunoglobulin (Ig) A acts as a first line of defense against respiratory pathogens. Mucosal IgA in salivary and nasal passages has a rapid response to antigens and can play a protective role against reinfection. The mainstay for analyzing SARS-CoV-2 infection and vaccine efficacy has been assessment of serum IgG levels; however, validated assays for assessment of mucosal IgA in clinical samples are necessary as new and adapted measures are generated to combat immune-evasive viral variants. A mucosal IgA assay was developed and tested through assessment of IgA levels in salivary samples from participants of the 2019nCoV-314/ NCT05973006 study. These participants had previously received ≥2 mRNA-based COVID-19 vaccinations prior to enrollment and received a single intramuscular study dose of NVX-CoV2601 (XBB.1.5) or bivalent vaccine (NVX-CoV2601 + NVX-CoV2373 [Wuhan]). Salivary samples were collected prior to vaccination on day 0 and on day 28 to assess response post vaccination. Both vaccine groups elicited a significant increase in anti–SARS-CoV-2 spike IgA against XBB.1.5. Furthermore, cross-reactivity via identification of anti-JN.1 and anti-Wuhan IgA was also observed. The detection of IgA in clinical mucosal samples through this assay will be a valuable tool in supporting vaccine development.

## INTRODUCTION

Immunoglobulin A (IgA) is the most predominant antibody class.^1^ It is present in serum but is most prevalent in mucosal tissues/secretions, such as the upper respiratory tract, nasal cavity, and saliva, in the form of secretory IgA (sIgA).^1^ Localized in the airways, IgA plays a key role in initial immune responses against inhaled antigens at the site of their entry into the body.^1^ Individuals with impaired IgA production/responses have been shown to have increased susceptibility to respiratory infection and recurrence, as well as development of chronic respiratory disease.^2^ Furthermore, sIgA is more resistant to some proteases than IgG and serum-derived IgA^1^, supporting its critical role in immune defense of the upper respiratory tract.^3^

IgA responses to SARS-CoV-2 infection are rapid, with detection of spike-specific IgA in serum within 2–5 days of symptom onset and a higher proportion of patients eliciting an early IgA response compared with an early IgG response.^4^ The rapid production of IgA preceding that of IgG^5^ makes it an ideal candidate for surveillance and early detection of infection; although there is a faster reduction in serum IgA than serum IgG levels.^4^ Notably, spike-specific mucosal IgA has been shown to persist after receipt of a COVID-19 vaccine primary series and additional doses^6^; however, one study reported that salivary IgA level increases observed after a first dose of an mRNA-based COVID-19 vaccine dropped to near baseline after 3 months and were recovered in only ∼30% of individuals after a second dose.^7^

The protective impact of IgA was also investigated in previously vaccinated healthcare workers who had also experienced at least one SARS-CoV-2 infection.^6^ In that study, those with breakthrough infections had significantly lower post-vaccination anti-spike serum IgA than those who were uninfected; notably, there were only modest differences in serum IgG levels between the infected and uninfected groups. Overall, nasal and salivary anti-spike SARS-CoV-2 IgA levels were higher than those for IgG. Nasal and salivary IgA levels correlated with one another; although, IgA levels were higher in nasal versus salivary secretions.

Despite the presence of anti-spike IgA in serum and mucosal secretions, IgG remains a primary measure for assessing immunogenicity of COVID-19 vaccines and testing for SARS-CoV-2 infection.^8^ IgA has protective potential by limiting viral spread and shedding, as reported for influenza in animal and human studies^9^; however, larger studies for influenza and other respiratory viruses are needed. In contrast to serological IgG, validated IgA assays for COVID-19 vaccines are limited in availability and clinical use^10,11^; although, as clinical investigations of mucosal IgA move forward, validated assays are needed.

The 2019nCoV-314/NCT05973006 study demonstrated the safety and immunogenicity of monovalent (NVX-CoV2601 [XBB.1.5]) and bivalent (NVX-CoV2601 + NVX-CoV2373 [Wuhan]) recombinant spike protein COVID-19 nanoparticle vaccines (with Matrix-M™ adjuvant) to ancestral SARS-CoV-2 and the XBB.1.5 strain in previously vaccinated adolescent participants.^12^ An exploratory endpoint of this study was to investigate the salivary anti-spike IgA responses within each of these vaccine groups; therefore, a mucosal IgA assay was developed, validated, and used for clinical testing.

## Methods

In the 2019nCoV-314 study, participants had previously received ≥2 mRNA-based COVID-19 vaccinations, followed by a single study dose of NVX-CoV2601 or bivalent vaccine.^13^ The trial protocol was approved by the Advarra Institutional Review Board and Independent Ethics Committee; the study performed in accordance with the Declaration of Helsinki and the International Conference on Harmonization Good Clinical Practice guidelines; and consent was provided by all participants.

Clinical saliva samples from adolescent participants of the 2019nCoV-314 study were collected on days 0 and 28 via the Salivette^®^ method (via an absorbent roll in the mouth), followed by isolation and freezing of saliva for storage. Total IgA (dimer and monomer) was measured via quantitative indirect ELISA against antibodies binding to SARS-CoV-2 recombinant spike protein (including XBB.1.5) as a reference standard to allow concentration measurement in ng/mL. Quality control (QC) samples were used in each run of the assay, comprised of high (IgA-spiked) QC, mid QC, and low QC saliva samples. The assay was validated based on the following parameters: precision, linearity, selectivity, specificity, sensitivity, robustness, and sample stability (to be described in detail in a separate publication). A set of pooled saliva samples tested for anti-spike IgA activity against XBB.1.5 was also tested for cross-reactivity to ancestral SARS-CoV-2 (Wuhan) and JN.1. Samples were pooled across participants based on similar IgA concentrations to generate sufficient volume for this analysis.

A Wilcoxon signed rank test was performed to compare mean concentrations of IgA at baseline with those on day 28 after study vaccination, and *p-*values were determined comparing day 0 and day 28 outcomes for each vaccine group. Seroresponse rates (SRRs) were calculated based on a >2-fold or >3-fold increase in post-vaccination titer from baseline (or from the lower limit of quantification if the baseline value was below this limit). SRR 95% CIs were calculated based on the Clopper–Pearson exact method.

## Results

Matched day 0 and day 28 samples were available for this analysis from a total of 347 participants in the 2019nCoV-314 study (NVX-CoV2601, N = 171; bivalent vaccine, N = 176). Anti-spike IgA titers (95% CI) against XBB.1.5 increased from 6.34 ng/mL (5.48–7.33) to 9.03 (7.74–10.55) in the NVX-CoV2601 group and from 7.03 (6.07–8.15) to 9.32 (7.99–10.85) in the bivalent vaccine group (**Figure 1**). Mucosal IgA responses were also observed regardless of participant baseline SARS-CoV-2 serostatus; however, the baseline seronegative population was small (n = 19) compared with the seropositive population (n = 328).

**Figure 1.**
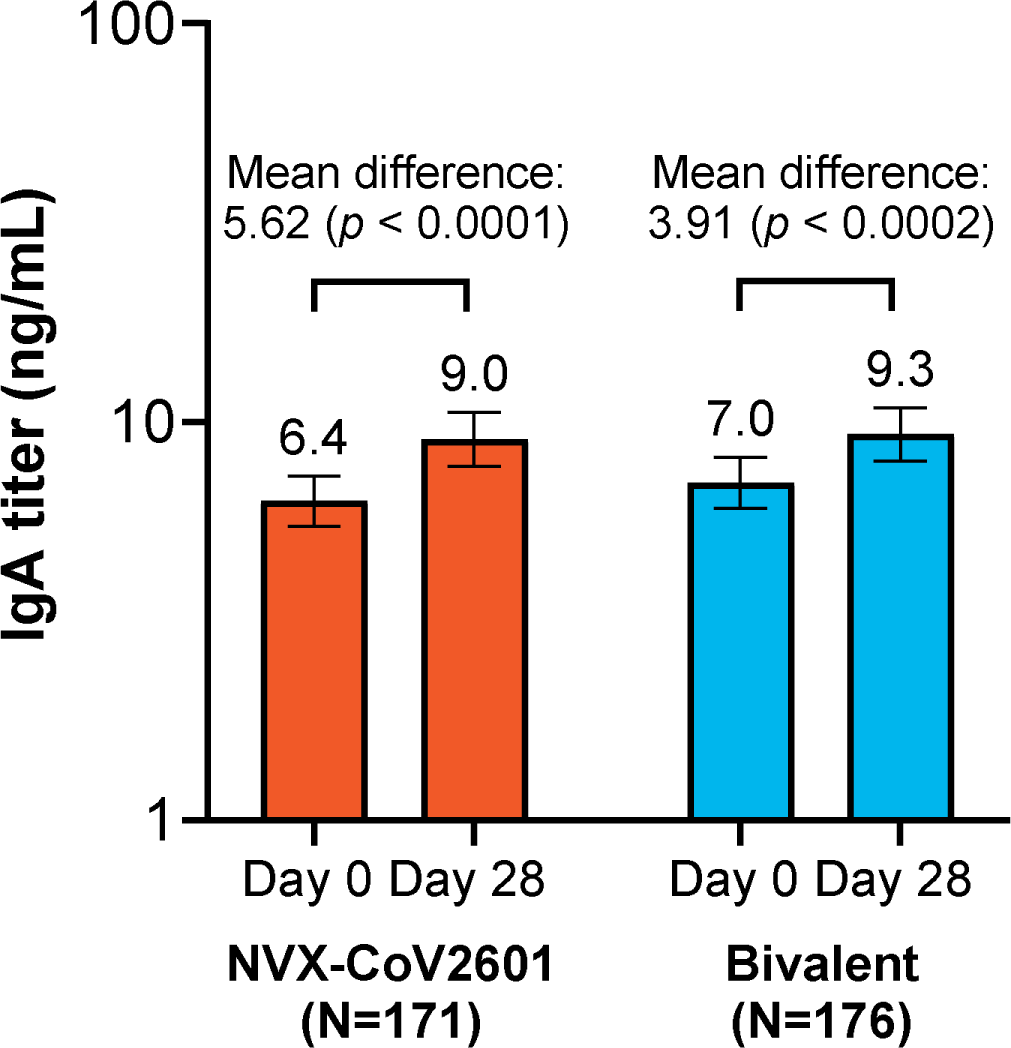
Anti-spike salivary IgA levels against SARS-CoV-2, variant XBB.1.5, from clinical samples from the 2019nCoV-314 study. IgA titers (ng/mL) at day 0 and day 28 after vaccination with NVX-CoV2601 or bivalent vaccine. IgA, immunoglobulin A.

SRRs to XBB.1.5 were calculated based on a 2-or 3-fold increase from baseline titer (**Table**). The SRR was 13% in the NVX-CoV2601 group with the 3-fold cutoff, which increased to 21% with the 2-fold cutoff. There were only slightly lower SRRs in the bivalent vaccine group: 7% and 18% at the 3-fold and 2-fold cutoffs, respectively. Seroresponse was observed regardless of baseline SARS-CoV-2 serostatus; however, SRRs in the seropositive subgroup trended similar to the overall population, whereas SRRs in the seronegative group were slightly lower.

**Table.** IgA seroresponse rates (SRRs) to anti-rS XBB.1.5 overall and by baseline SARS-CoV-2 serostatus from saliva samples of participants in the 2019nCoV-314 study.

|  | Overall |  | Baseline seropositive |  | Baseline seronegative |  |
| --- | --- | --- | --- | --- | --- | --- |
| **SRR, n (%)<sup>a</sup>** | **NVX-CoV2601**<br>**(N = 171)** | **Bivalent**<br>**(N = 176)** | **NVX-CoV2601**<br>**(n = 159)** | **Bivalent**<br>**(n = 169)** | **NVX-CoV2601**<br>**(n = 12)** | **Bivalent**<br>**(n = 7)** |
| >3-Fold rise | 22 (12.9) | 13 (7.4) | 21 (13.2) | 13 (7.7) | 1 (8.3) | 0 |
| >2-Fold rise | 35 (20.5) | 32 (18.2) | 33 (20.8) | 31 (18.3) | 2 (16.7) | 1 (14.3) |

<sup>a</sup>SRRs were calculated based on a >2-fold or >3-fold increase in post-vaccination titer from baseline (or from the lower limit of quantification if the baseline value was below this limit).
Abbreviations: IgA, immunoglobulin A; rS, recombinant Spike; SRR, seroresponse rate.

Reactivity of pooled samples from the 2019nCoV-314 study to XBB.1.5 was assessed alongside cross-reactivity to the JN.1 strain and ancestral SARS-CoV-2 (Wuhan) (**Figure 2**). Against XBB.1.5,11/11 pooled samples had a mean concentration of IgA greater than that for the low QC samples. Against JN.1, 8/11 samples had a mean concentration above the low QC; 11/11 samples were above the low QC for Wuhan.

**Figure 2.**
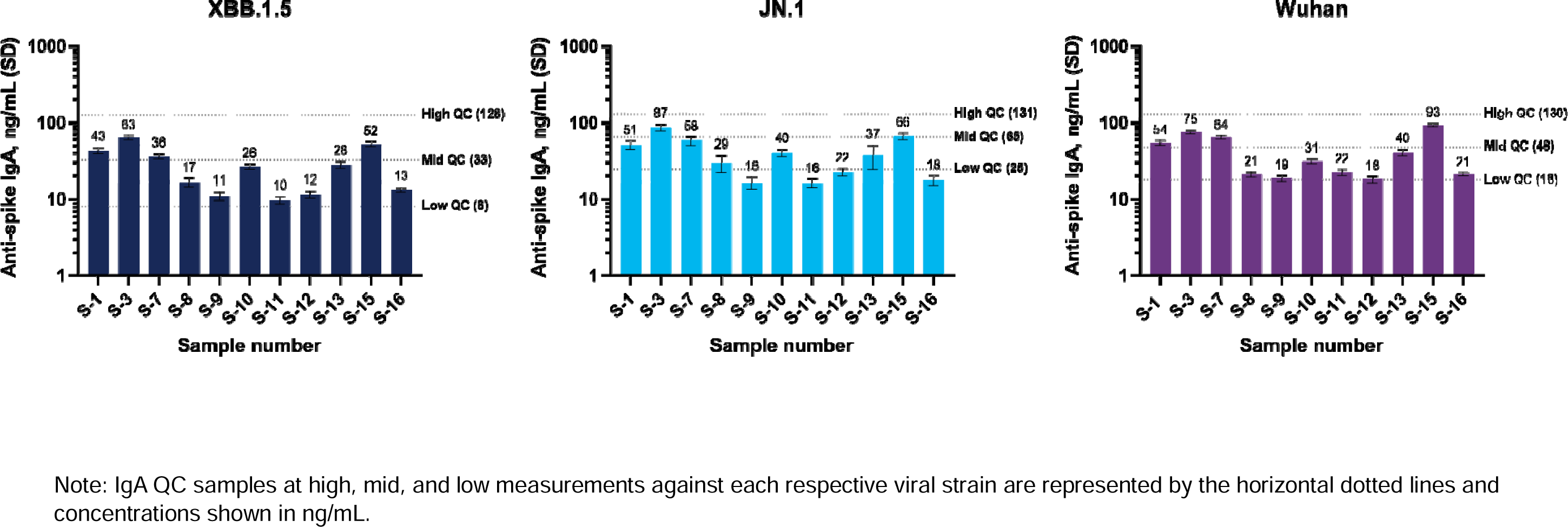
Cross-reactivity of salivary IgA after immunization with NVX-CoV2601 (XBB.1.5 vaccine) in pooled serum samples from the 2019nCoV-314 study. Anti-spike IgA concentrations against SARS CoV-2, variant strains XBB.1.5 or JN.1, or Wuhan SARS-CoV-2 at 28 days after vaccination with NVX-CoV2601 in pooled salivary samples from the 2019nCoV-314 study. Samples displayed are those that had sufficient sample available to be tested across all three strains. GMT, geometric mean titer; IgA, immunoglobulin A; S, sample; SD, standard deviation; QC, quality control.

## Discussion

Proof of concept for this salivary IgA assay was established based on a significant increase in anti–SARS-CoV-2 spike IgA against XBB.1.5 in clinical saliva samples 28 days after study vaccination. Not only were matched spike (XBB.1.5) responses observed but there were also cross-reactivity and detection of anti-JN.1 and anti-Wuhan IgA. Notably, an increase in IgA from baseline was detected at day 28, even though these responses were known to be rapid^4^ and might have peaked at an earlier timepoint.

In our analysis, a subset of 11 pooled samples (pooled based on similar IgA concentrations) each had a mean IgA concentration against XBB.1.5 that was greater than the low QC samples. Similarly, a separate study of triple-vaccinated healthcare workers (the majority having received mRNA-based COVID-19 vaccines), elicited mucosal IgA responses specific to wild-type SARS-CoV-2 at 5 weeks post 3^rd^ dose.^14^ In that study, spike-specific mucosal IgA levels were higher in individuals with prior SARS-CoV-2 infection, which is in line with the higher SRRs in baseline seropositive versus seronegative participants observed here and reported in the Table.

The majority (93%) of participants from whom samples were obtained for this assessment were baseline SARS-CoV-2 seropositive, which is representative of the global population.^15^ The prevalence of SARS-CoV-2 infection and COVID-19, with almost 300,000 global cases documented in September 2024,^16^ highlights a need for new and improved methods of identifying infected individuals and reducing infection and transmission. IgA has been found to have more potent neutralization potential than IgG for early SARS-CoV-2 infection^17^ and mucosal IgA levels (but not necessarily IgG) may align with protection from breakthrough omicron infection^14^. Continued subsequent doses of current mRNA-based COVID-19 vaccines indicate that additional boosting of mucosal IgA responses is lacking.^18^

The data presented here, and the importance of mucosal immunity, underscore the need for more in-depth examination of mucosal IgA to develop mechanisms to harness its strengths, improve immune responses, provide both long-term protection and cross-reactivity,^2^ and define levels for correlates of protection across COVID-19 vaccines. An example where the benefits of mucosal IgA response has been employed are aerosolized intranasal vaccines. These vaccines induce a localized IgA response at the site of pathogen entry to enhance mucosal immunity; additionally, they have feasibility of use and economic advantages over injectable formulations.^19^ Initial investigations indicate that intranasal COVID-19 vaccines can elicit a mucosal IgA response to the matched viral strain and cross-reactivity to related variants.^20^ Overall, the outcomes from assessment of this assay, in addition to the totality of reports of mucosal IgA responses after vaccination, should provide initial guidance and confidence to conduct future IgA clinical studies.

There are some limitations to this proof-of-concept assessment. First, the clinical sample population was comprised of adolescents only, and there may be differences in response based on age, comorbidity, concomitant medication, or another unique factor that would be present in a broader-age population. Also, based on the small sample size, no formal conclusions can be drawn regarding the clinical significance of these findings, and additional analyses are warranted to properly evaluate the production of SARS-CoV-2 spike-specific IgA and how it may relate to the development of immunity. Mucosal IgA was detected in saliva samples collected from individuals who received intramuscular Novavax COVID-19 vaccine; however, this may not reflect potentially higher levels that could occur after intranasal vaccine delivery.

As cases of COVID-19 remain prevalent, investigations into new ways to protect from SARS-CoV-2 infection must continue. The detection of IgA in clinical saliva samples will be a useful tool in developing vaccines that aim to elicit and improve mucosal immunity and break-in on the cycle of infection through increased protection and decreased transmission.

## AUTHOR CONTRIBUTIONS

Chijioke Bennett and Raburn M. Mallory were involved in the study design. Mingzhu Zhu, Edmond Massuda, Urvashi Patel, Raj Kalkeri, Shane Cloney-Clark, and Joyce S. Plested verified the data. Gordon Chau performed the statistical analyses. All authors had access to the data and were involved in data interpretation, manuscript preparation, and review. All authors provided approval to submit for publication.

## ACKNOWLEDGMENTS

We thank all the study participants who volunteered for this study. This study was funded by Novavax, Inc. Medical writing and editorial support were provided by Kelly M. Fahrbach, PhD, CMPP, and Ebenezer M. Awuah-Yeboah, BS, of Ashfield MedComms (New York, USA), an Inizio company, supported by Novavax, Inc.

## DATA AVAILABILITY STATEMENT

The information and data that support the findings of the study are available at https://clinicaltrials.gov/study/NCT05973006. Requests submitted to the corresponding author will be considered upon publication of this article, and deidentified participant data may be provided.

## ETHICS STATEMENT

The trial protocol was approved by the Advarra Institutional Review Board and Independent Ethics Committee, the study performed in accordance with the Declaration of Helsinki and the International Conference on Harmonization Good Clinical Practice guidelines, and consent was provided from all participants.

